# A novel two-hit insomnia and inflammation rodent model of depressive-like behaviors

**DOI:** 10.1101/2024.08.01.24311351

**Authors:** Junhua Mei, Xinhua Song, Ying Wang, Honggang Lyu, Guang Wang, Chao Chen, Honghan Zhang, Chao Wang, Xin-hui Xie, Guohua Chen, Zhongchun Liu

**Author notes:** Co-first author (Junhua Mei, Xinhua Song, Ying Wang, Honggang Lyu). Corresponding authors: 1. Zhongchun Liu., Address: 1) Department of Psychiatry, Renmin Hospital of Wuhan University, No. 99 Jiefang Road, Wuchang District, Wuhan, Hubei, PR China. ZIP: 430060. 2) Taikang center for life and medical sciences, Wuhan University, Wuhan, PR China. ZIP: 430071., 2. Guohua Chen, Address: Department of Neurology, Wuhan First Hospital, No. 215 Zhongshan Road, Wuhan 430022, China., 3. Xin-hui Xie, Address: Department of Psychiatry, Renmin Hospital of Wuhan University, No. 99 Jiefang Road, Wuchang District, Wuhan, Hubei, PR China. ZIP: 430060.

## Abstract

**Background:** Systemic inflammation and insomnia often co-occur in patients with depression. However, there is no suitable animal model to investigate the relationship between inflammation, sleep deprivation (SD), and depression.

**Methods:** To model interactions between insomnia, inflammation, and depression, we developed a novel “two-hit” rodent model of depressive-like behaviors using continuous SD followed by daily lipopolysaccharide (LPS) treatment. Control groups received SD, LPS, or sterile phosphate-buffered salinealone. The model’s validity was assessed at the cellular and molecular levels, with fluoxetine rescue applied to confirm model validity.

**Results:** The model group demonstrated significant depressive-like behaviors that were rescued by fluoxetine treatment. Transcriptomic analysis revealed alterations in neuroinflammation and synaptic plasticity pathways within the hippocampus and prefrontal cortex (PFC) of model rats. Western blotting validated alterations in key protein markers related to both processes, and immunofluorescence confirmed microglia and astrocyte activation, indicative of neuroinflammation. Additionally, transmission electron microscopy and Golgi-Cox staining revealed reduced synapse and dendritic spine density in the model group. Fluoxetine treatment reversed these structural changes. Sixteen genes associated with neuroinflammation and synaptic function were validated in human genetic studies by transcriptome-wide association analysis.

**Conclusion:** This reliable two-hit model will be useful for investigating the roles of insomnia and inflammation in depression.

## INTRODUCTION

Depression is extremely common and frequently recurs, and it is expected to be the leading disease in terms of global burden by 2030 [1]. Depression is a complex and clinically and biologically heterogeneous, with a multifactorial etiology [2]. Traditional hypotheses to explain depression, such as the neurotransmitter and receptor hypothesis, the hypothalamic-pituitary-adrenal axis hypothesis, and the neuroplasticity hypothesis, certainly provide insights into the etiology of depression. However, these hypotheses do not fully explain the underlying pathobiology nor do they completely explain the complex clinical phenotypes [3], and other mechanisms are now thought to contribute to the disease process. In this regard, and based on clinical observations, recent studies have focused on the roles played by insomnia and inflammation in depression [4–6].

The relationship between insomnia, inflammation, and depression is complex. Insomnia is commonly observed in individuals with depression [7], and insomnia may be a predictor of depression recurrence [8]. A meta-analysis of individuals with insomnia concluded that insomnia was associated with elevated systemic inflammatory markers [9]. Insomnia also activates multiple inflammatory pathways, such as the transcription of proinflammatory nuclear factor kappa-light-chain-enhancer of activated B cells (NF-κB) pathways and proinflammatory interleukin-6 (IL-6) [10, 11]. Similarly, there is a well-established relationship between inflammation and depression. Several meta-analyses have demonstrated that, even after accounting for confounding, inflammatory markers are significantly increased in patients with depressive disorders compared with healthy controls [12, 13]. Complementing these cross-sectional data, longitudinal cohort studies have also indicated that children with elevated serum IL-6 and C-reactive protein were at significantly increased risk of developing depressive disorders during adolescence [14].

Importantly, inflammation and insomnia often co-occur in patients with depression, and insomnia-related and inflammatory processes induced by infection and psychological stress elevate depression risk [15, 16]. Clinical studies provide further evidence of an association between inflammation and the severity of insomnia or depressive symptoms [6]. Moreover, mediation analyses of the large National Health and Nutrition Examination Survey (NHANES) community sample also revealed that increased inflammatory markers might mediate the association between insomnia and depression [5]. Hence, Irwin and Piber proposed a two-hit hypothesis of depression, in which insomnia and inflammation collaborate as “two hits” in a population especially vulnerable to developing depression [16]. Despite this abundance of clinical evidence and a plausible hypothesis, there is a conspicuous absence of animal models to rigorously investigate the underlying biological mechanisms. Therefore, here we propose a novel rodent model of depression-like symptoms based on the two-hit hypothesis.

Our model simulates insomnia and inflammation using sleep deprivation (SD) and lipopolysaccharide (LPS) treatment, respectively. We evaluate our model from the etiological, phenomenological, and therapeutic perspectives. The induction of depressive-like behaviors by combined insomnia and inflammation was confirmed through behavioral assessments. Transcriptomic analysis defined the molecular signaling effects of the two hits of SD and inflammation on the hippocampus and prefrontal cortex (PFC), with further confirmatory exploration of the underlying pathobiology with western blotting, immunofluorescence, transmission electron microscopy (TEM), and Golgi-Cox staining validating the transcriptional patterns. Furthermore, we performed fluoxetine rescue experiments to assess the efficacy of fluoxetine in this model. Finally, we provide genetic evidence from human transcriptome-wide association analysis (TWAS) that supports the two-hit model and implicates neuroinflammation and synaptic dysfunction in depression pathology.

## MATERIALS AND METHODS

### The two-hit depressive-like model

#### Animals

All experiments were conducted on eight-week-old male Sprague Dawley rats purchased from Liaoning Changsheng Biotechnology Co., Ltd. (Liaoning, China, License Number: SCXK (Liao) 2020-0001). Rats were acclimatized to the new surroundings for seven days before commencing experiments. The Animal Ethics and Welfare Committee at Renmin Hospital of Wuhan University approved all experimental protocols (WDRM Dong (Fu), No. 20230603C).

#### Sleep deprivation

Chronic SD was conducted using an improved multi-platform water environment method [17]. Rats were housed each day in 75 x 50 x 36 cm^3^ SD boxes containing 8 cm diameter, 8 cm high platforms spaced 15 cm apart, with water injected to 1 cm above the platforms. Rats underwent continuous SD for 18 days, with each day lasting 18 hours (15:00 – 09:00). The water temperature during SD was maintained at 25℃ ± 2℃.

#### LPS treatment

LPS (O111:B4, Sigma-Aldrich, St. Louis, MO) was administered intraperitoneally (i.p.) in sterile phosphate buffered saline (PBS) at a dose of 0.8 mg/kg/day, calculated as a volume of 5.0 ml/kg.

#### Fluoxetine treatment

Fluoxetine (J20170022, Patheon, Bourgoin-Jallieu, France) was dissolved in drinking water and administered by oral gavage at a dose of 10 mg/kg/day in a volume of 2 mg/ml.

#### Model experiment

Forty-eight Sprague Dawley rats were acclimatized for seven days. Then, after the first behavioral assessment (sucrose preference test, SPT), rats were randomly assigned to four groups: control (CON, n = 12), LPS treatment (LPS, n = 12), sleep deprivation (SD, n = 12), and the model group (MOD, n = 12). Subsequent procedures are shown in **Fig. 1A**. The CON group was housed normally in cages and received a daily i.p. injection of 5.0 ml/kg sterile PBS solution from days 18 to 21. The LPS group was housed normally and received a daily i.p. injection of 0.8 mg/kg LPS solution from days 18 to 21. The SD group underwent 18 hours of SD daily from days 1 to 18, followed by a daily i.p. injection of 5.0 ml/kg sterile PBS solution from days 18 to 21. The MOD group underwent 18-hour SD daily from days 1 to 18, followed by a daily i.p. injection of 0.8 mg/kg LPS solution from days 18 to 21.

**Fig. 1.**
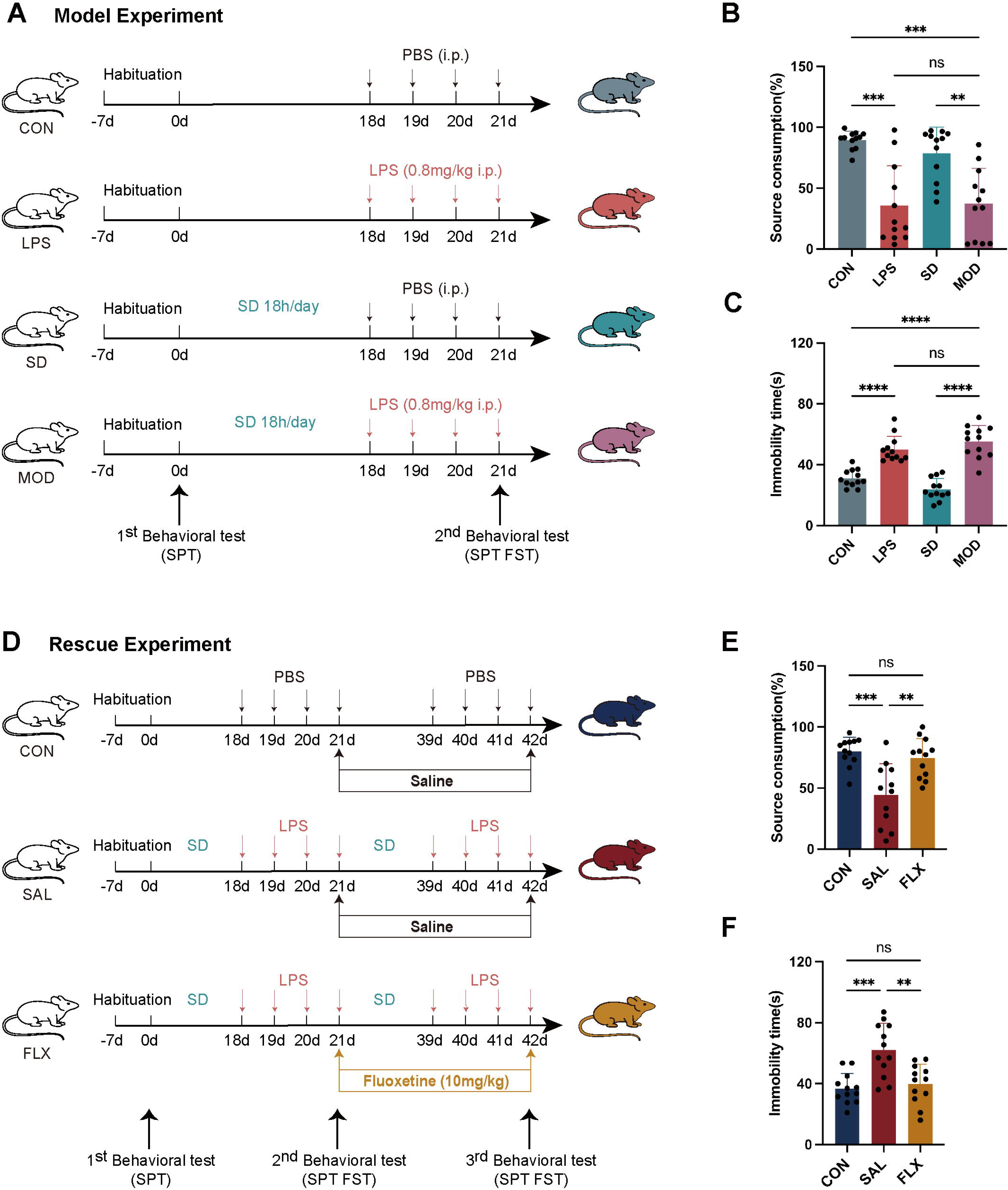
Experimental paradigm and behavioral test outcomes in SD– and LPS-treated rats. Schematics of the model (A) and rescue experiment (D) design. Source consumption preference in the SPT (n = 12) (B, E). Immobility times in the FST (n = 12) (C, F). Data are expressed as mean ± SD. \**p* < 0.05, \*\**p* < 0.01, \*\*\**p* < 0.001, \*\*\*\**p* < 0.0001 by one-way ANOVA with Fisher’s LSD multiple-comparisons test.

On the 21^st^ day, a second set of behavioral assessments were conducted (the SPT and the forced swimming test (FST)) to evaluate depressive-like behaviors in each group. After the FST, each group was euthanized and hippocampal and PFC tissue excised.

#### Fluoxetine rescue experiment

The flowchart for the fluoxetine rescue experiment is shown in **Fig. 1D**. Following a seven-day acclimatization period, 36 rats were randomly assigned to two groups at a 1:2 ratio: the control group (CON, n = 12) and the model group (MOD, n = 24). Subsequent procedures were similar to those in the model experiment. On the 22^nd^ day, after the second behavioral assessment, the model group was randomly divided into two subgroups: the physiological saline treatment group (SAL, n = 12) and the fluoxetine treatment group (FLX, n = 12).

Fluoxetine (10 mg/kg) was administered daily via gastric lavage to the fluoxetine group, while the saline group received an equal volume of physiological saline daily. The control group was maintained under normal cage conditions. The rats underwent the same 21-day modeling as described in the model experiment. On the 42^nd^ day, the third set of behavioral assessments was conducted, including the SPT and FST, to evaluate depressive-like behaviors in each group. Then, all rats were euthanized, and hippocampus and PFC tissue were excised.

#### Behavioral assessments

Details of the behavioral assessments (SPT, FST) are provided in **Supplemental File 1**.

### Transcriptome analysis

#### Principal component analysis for comparative analysis

Principal component analysis (PCA) was performed to examine the differences between samples [18].

#### Clustering and visualization of gene expression

The R package “*ClusterGVis*” was used to perform *k*-means clustering and to visualize differences in gene expression profiles from the hippocampus and PFC between the different groups (https://github.com/junjunlab/ClusterGVis).

#### Differentially-expressed genes (DEGs) and enrichment analysis

Differential expression analysis (DEA) was performed using the “*DESeq2*” package [19] in R to identify DEGs between the experimental groups (MOD, LPS, and SD) and the control group. Genes with an absolute fold change ≥ 1.2 and an adjusted *p*-value (adj. *p*-value) < 0.05 were considered statistically significant DEGs. To further investigate molecular mechanisms in the two-hit model, gene set enrichment analysis (GSEA) and Gene Ontology (GO) and Kyoto Encyclopedia of Genes and Genomes (KEGG) analyses were performed to explore signaling pathways in both the hippocampus and PFC using the R package “*clusterProfiler*” [20]. Next, as an exploratory analysis, Bayesian networks inferred from gene expression data based on enrichment analysis results were visualized using the “*CBNplot*” package in R [21].

### Validation of depression-related biological phenotypes

We used a battery of techniques to examine depression-related biological phenotypes in the hippocampus and PFC related to the transcriptomic results. Fiji/ImageJ software was used to analyze western blotting, immunofluorescence, TEM, and Golgi-Cox staining images. The specific methods are detailed in **Supplemental File 1**.

#### Western blotting

Western blotting was used to confirm changes in expression of synaptic plasticity (PSD95, SYN-1, SYP, and BDNF) and neuroinflammation (NLRP3, ASC) markers. The following primary antibodies were used: PSD95 (ABclonal, Woburn, MA; A7889, 1:2000), SYN-1 (Proteintech, Rosemont, IL; 20258-1-AP, 1:4000), SYP (Cell Signaling Technology, Danvers, MA; 5461T, 1:2000), BDNF (Abcam, Cambridge, UK; ab108319, 1:2000), NLRP3 (Abcam, ab263899, 1:1000), ASC (ABclonal, A1170, 1:1000).

#### Immunofluorescence

Immunofluorescence was used to detect and visualize specific proteins or antigens in cells or tissues. The immunofluorescence method is detailed in **Supplemental File 1**.

#### Transmission electron microscopy

TEM is a powerful technique used to visualize the ultrastructure of biological specimens at near-atomic resolution. For accurate quantification, we differentiated synaptic structures according to the following criteria: both pre– and postsynaptic thickened structures and at least three vesicles within a 0.1 µm range on the presynaptic side. We quantified the number of synapses (measured in micrometers squared, µm^2^), the area of readily released vesicle pools, and the dimensions of the postsynaptic densities to observe synaptic ultra-microstructures in the hippocampus [22].

#### Golgi-Cox staining

Golgi-Cox staining was used to visualize neurons and their dendritic trees in hippocampal tissue sections (**Supplemental File 1**). Dendrites on the secondary branches of neurons were selected, with the length of each dendritic segment exceeding 10 µm.

### Transcriptome-wide association studies

To further validate whether DEGs from the two-hit model are also associated with human depression, we used the FUSION (http://gusevlab.org/projects/fusion/) framework to perform TWAS [23]. This method leverages tissue-specific expression quantitative loci (eQTL) information across multiple human tissues to identify putative causal genes for a phenotype. Here, we performed brain-specific tissue TWAS leveraging 13 brain tissues available from GTExV8 (https://gtexportal.org/home/aboutAdultGtex). The depression Genome-wide association study (GWAS) summary data was downloaded from the iPSYCH website (https://ipsych.dk/en/research/downloads). A false discovery rate (*FDR.p*) was applied in multiple testing.

## RESULTS

### Behavioral assessments

The model group displayed significant depressive-like behaviors (**Fig. 1B,C**). Compared to controls, model rats exhibited a significant decrease in SPT (*p* < 0.01) and a significant increase in immobility time in the FST (*p* < 0.01). The LPS group also displayed similar behavioral alterations. By contrast, the SD group did not show significant depressive-like behaviors.

In the fluoxetine rescue experiment, fluoxetine rescued these depressive-like behaviors induced by combined SD and LPS (**Fig. 1E,F**).

### Hippocampus and PFC transcriptomes

#### PCA

PCA plots depicting the distribution of samples based on gene expression profiles in the hippocampus and PFC are shown in **Supplemental Fig. 1**.

#### Clustering and visualization of gene expression

Analysis of hippocampus and PFC transcriptomes identified eight distinct clusters (C1–C8) (**Fig. 2A,B**). In the hippocampus, cluster C5 was enriched for inflammation and immune pathways, including those involved in leukocyte and neutrophil migration and regulation of immune effector processes. Additionally, cluster C1 revealed contributions from cognition and learning or memory processes in the model group.

**Fig. 2.**
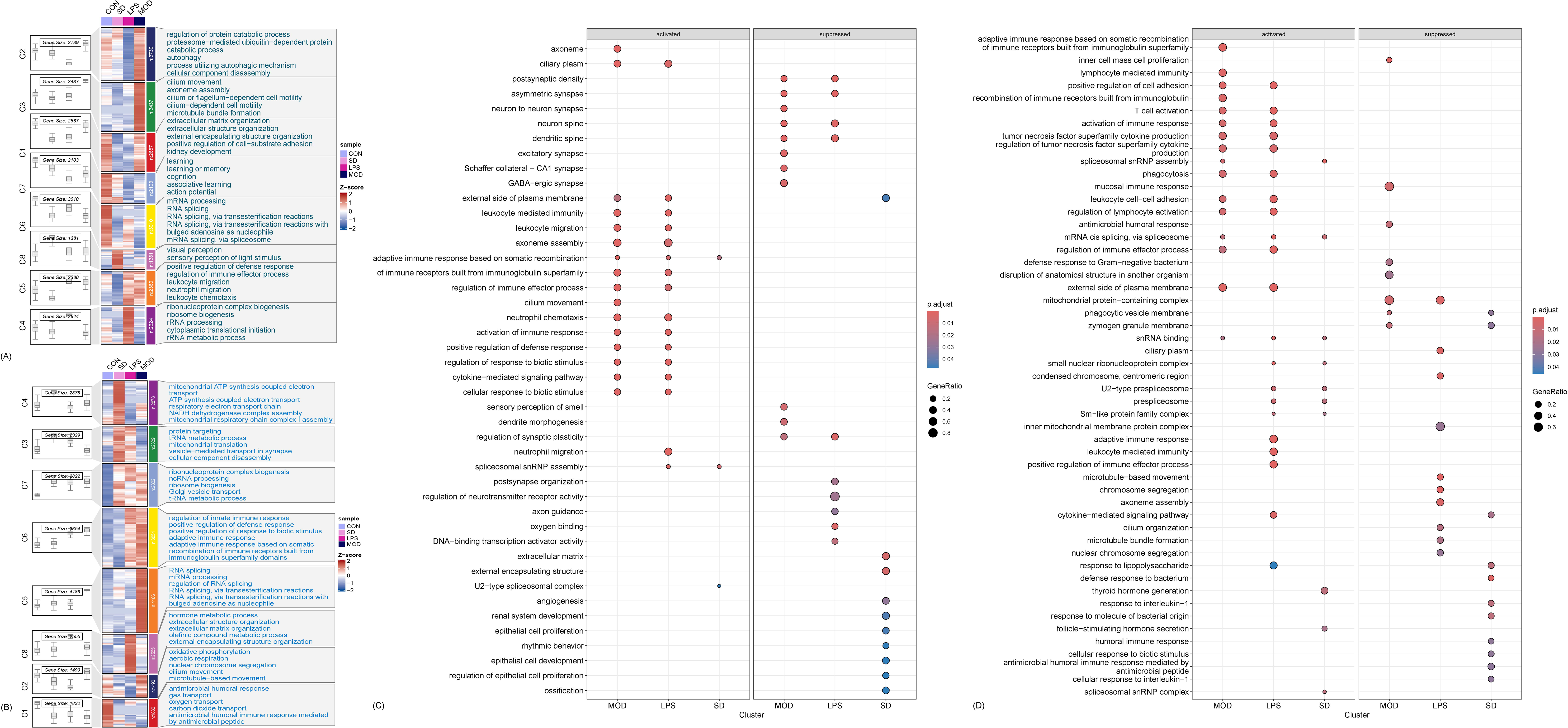
Gene expression heatmaps and dotplots of enrichment analysis. Cluster analysis was conducted on the gene expression matrix from the hippocampus (A) and PFC (B). Enrichment analysis was performed for each cluster, and boxplots and heatmaps were generated to display the expression levels of each group. The GSEA results of the experimental groups from the hippocampus (C) and PFC (D) demonstrate differences in enriched pathways between groups.

In the PFC, cluster C6 highlighted pathways associated with inflammatory and immune responses, while there was significant expression of synapse-related pathways, including vesicle-mediated transport in synapses, in cluster C3 (**Fig. 2B**).

#### DEGs and enrichment analysis

In the hippocampus, 238 DEGs were identified in the model group, of which 195 were upregulated and 133 were downregulated. GSEA analysis revealed significant enrichment of inflammatory and immune response pathways in the model group, including those related to leukocyte migration, immune activation, and cytokine-mediated signaling. Conversely, there was downregulation of synaptic pathways involved in postsynaptic density, asymmetric synapses, neuron-to-neuron synapses, GABAergic synapses, and regulation of synaptic plasticity in the model group (**Fig. 2C**).

In the PFC, 123 DEGs were identified in the model group, of which 64 were upregulated and 59 were downregulated. Similarly, GSEA analysis also showed significant activation of inflammatory and immune response pathways (**Fig. 2D**). Detailed DEA results are presented in **Supplemental Table 1**.

Over-representation analysis (ORA) of DEGs within the GO database yielded similar results. In the model group, upregulated DEGs in the hippocampus were significantly enriched for inflammatory and immune response pathways, while downregulated DEGs were enriched for synapse-related pathways (**Fig. 3A,C**). Additionally, to provide a deeper understanding of the underlying mechanisms behind these molecular pathways (**Fig. 3B, D, F, and H**), we explored Bayesian networks among these pathways. Detailed ORA results are presented in **Supplemental Table 2**.

**Fig. 3.**
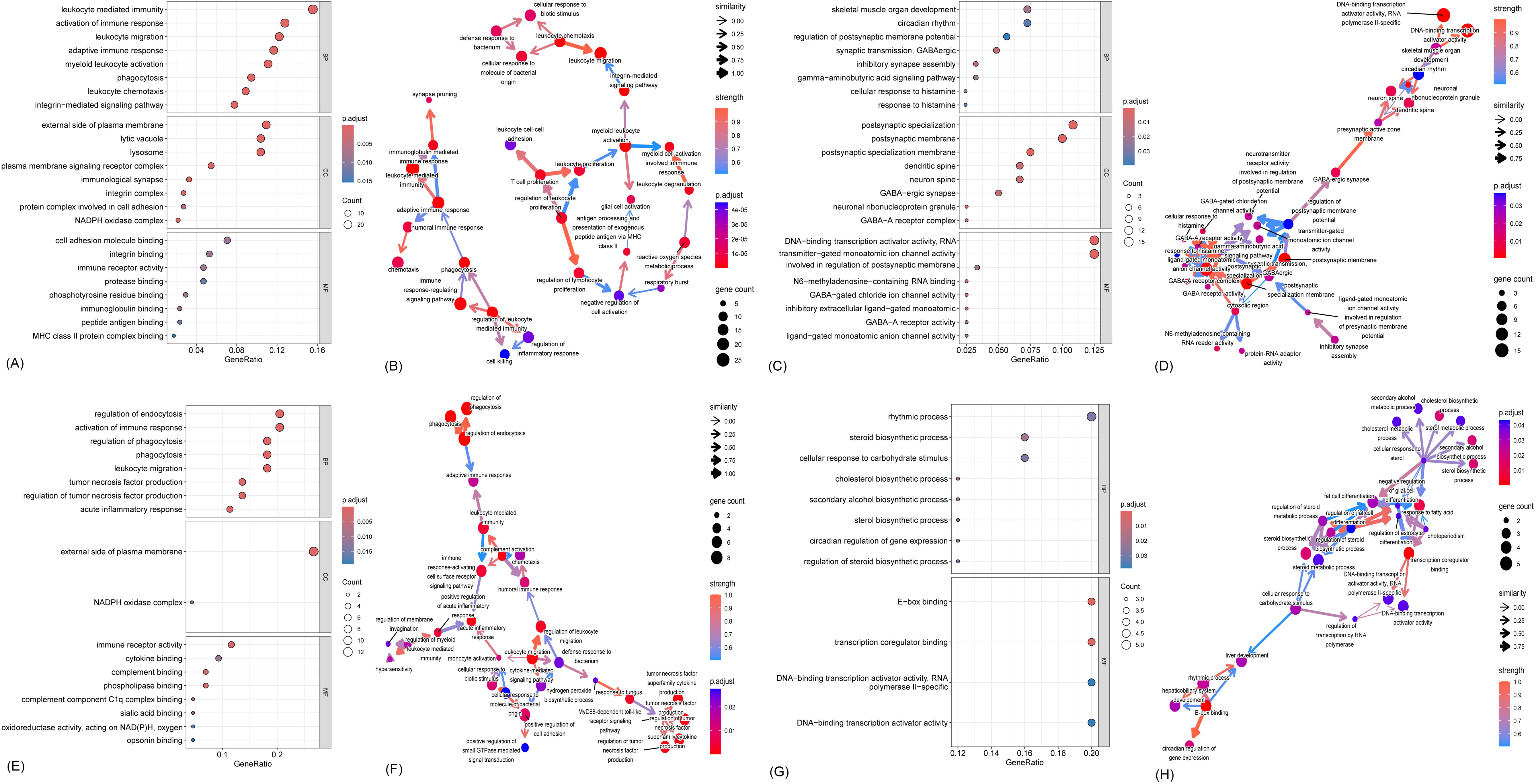
Enrichment analysis using the DEGs and Bayesian networks inferred from gene expression data in the hippocampus and PFC of the MOD group. (A and B) Results of the upregulated DEGs in the hippocampal MOD group, including Biological Processes (BP), Molecular Functions (MF), and Cellular Components (CC), and visualization of the interactions between these pathways. (C and D) Results of downregulated DEGs in the hippocampal MOD group. (E and F) Results of the upregulated DEGs in the PFC MOD group. (G and H) Results of the downregulated DEGs in the PFC MOD group.

### Validation of depression-related biological phenotypes

Transcriptomic analysis suggested involvement of neuroinflammation and synaptic plasticity in the model group. We therefore applied several methodologies to validate these alterations in neuroinflammation and synaptic processes within the hippocampus and PFC.

#### Neuroinflammation

As shown in **Fig. 4**, in the hippocampus and PFC of model rats, there was significant upregulation (*p* < 0.05) of inflammatory proteins such as NLRP3 and ASC compared with control. Immunofluorescence analysis confirmed cellular-level changes, as evidenced by increased GFAP^+^ and IBA-1^+^ cells, suggesting activation of astrocytes and microglia in the dentate gyrus (DG), CA1, and CA3 regions of the hippocampus. Similar observations were noted in the PFC.

**Fig. 4.**
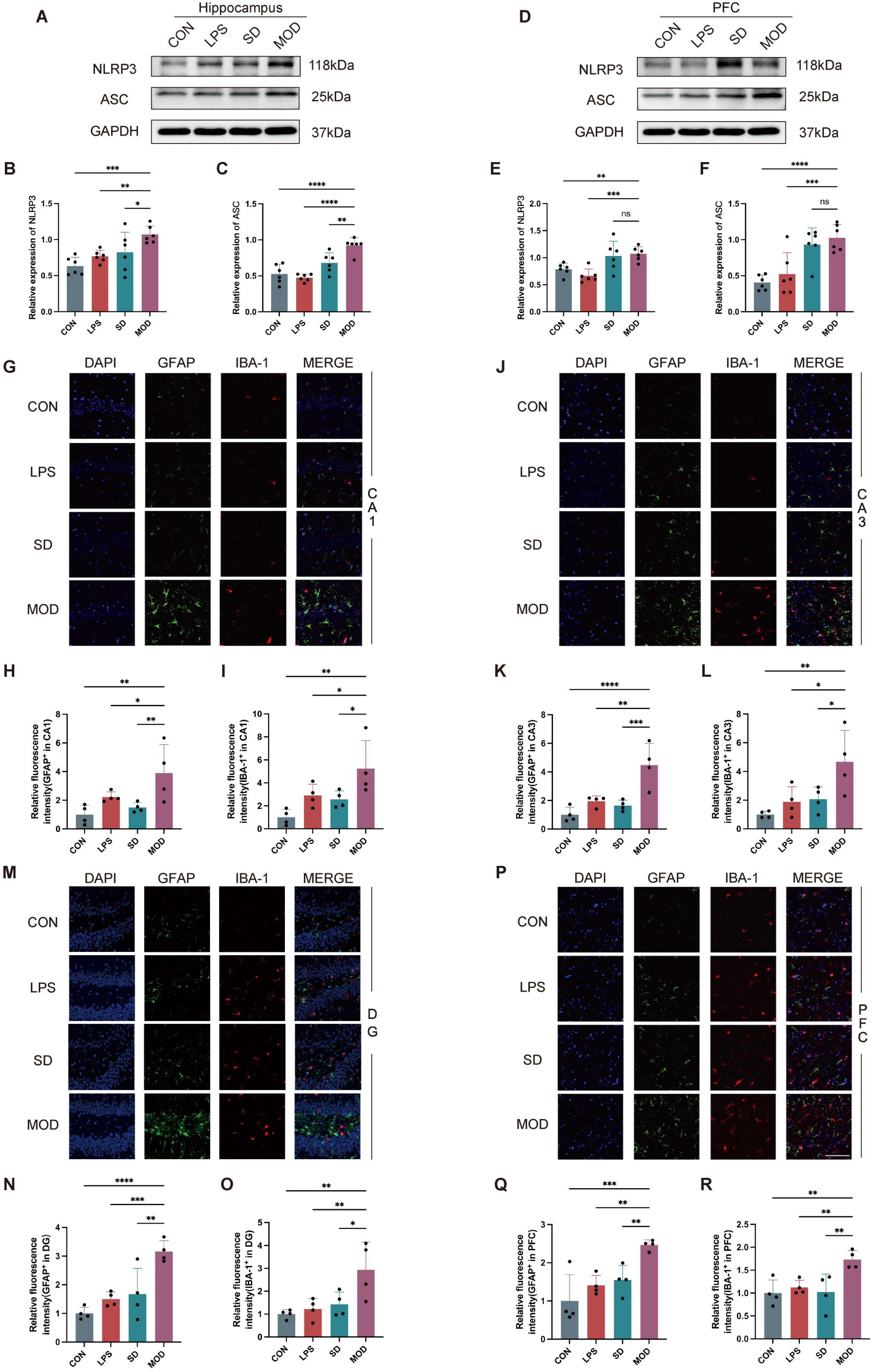
SD plus LPS treatment induces neuroinflammation in both the hippocampus and PFC. Western blotting analysis showing an increase of NLRP3 and ASC in MOD hippocampus (A) and PFC (D). Quantification of the NLRP3/GAPDH and ASC/GAPDH ratios (n = 6) in hippocampus (B, C) and PFC tissue (E, F). Representative immunofluorescence images of GFAP^+^ and IBA-1^+^ cells in CA1 (G), CA3 (J), DG (M), and PFC (P) in the indicated groups. Scale bars, 100 μm (white). Quantitation of GFAP^+^ cell relative fluorescence intensity in the indicated groups (H, K, N, Q). Quantitation of the IBA-1^+^ cell relative fluorescence intensity in the indicated groups (I, L, O, R). Data are expressed as mean ± SD. \**p* < 0.05, \*\**p* < 0.01, \*\*\**p* < 0.001, \*\*\*\**p* < 0.0001 by one-way ANOVA with Fisher’s LSD multiple-comparisons test.

In the fluoxetine rescue experiment, as expected, there was decreased expression of NLRP3 and ASC in both the hippocampus and PFC in the fluoxetine group (*p* < 0.05), and immunofluorescence analysis also revealed that fluoxetine rescued alterations in GFAP^+^ and IBA-1^+^ cells (**Supplemental Fig. 5**).

#### Synaptic plasticity

As shown in **Fig. 5**, decreased expression of synapse-related proteins, including PSD95, SYN, and SYP, was observed in the hippocampus and PFC of model rats (*p* < 0.05). Additionally, BDNF expression was significantly downregulated (*p* < 0.05). Moreover, combined SD and LPS treatment produced ultra-structural changes in synaptic structures. TEM revealed a decrease in synapse numbers and the average size of synaptic vesicles of the readily releasable pool (RRP), together with a reduction in the length and thickness of the postsynaptic density (PSD) in the hippocampus. Golgi-Cox staining indicated a reduction in the number of dendritic spines, which were shorter, narrower, and elongated.

**Fig. 5.**
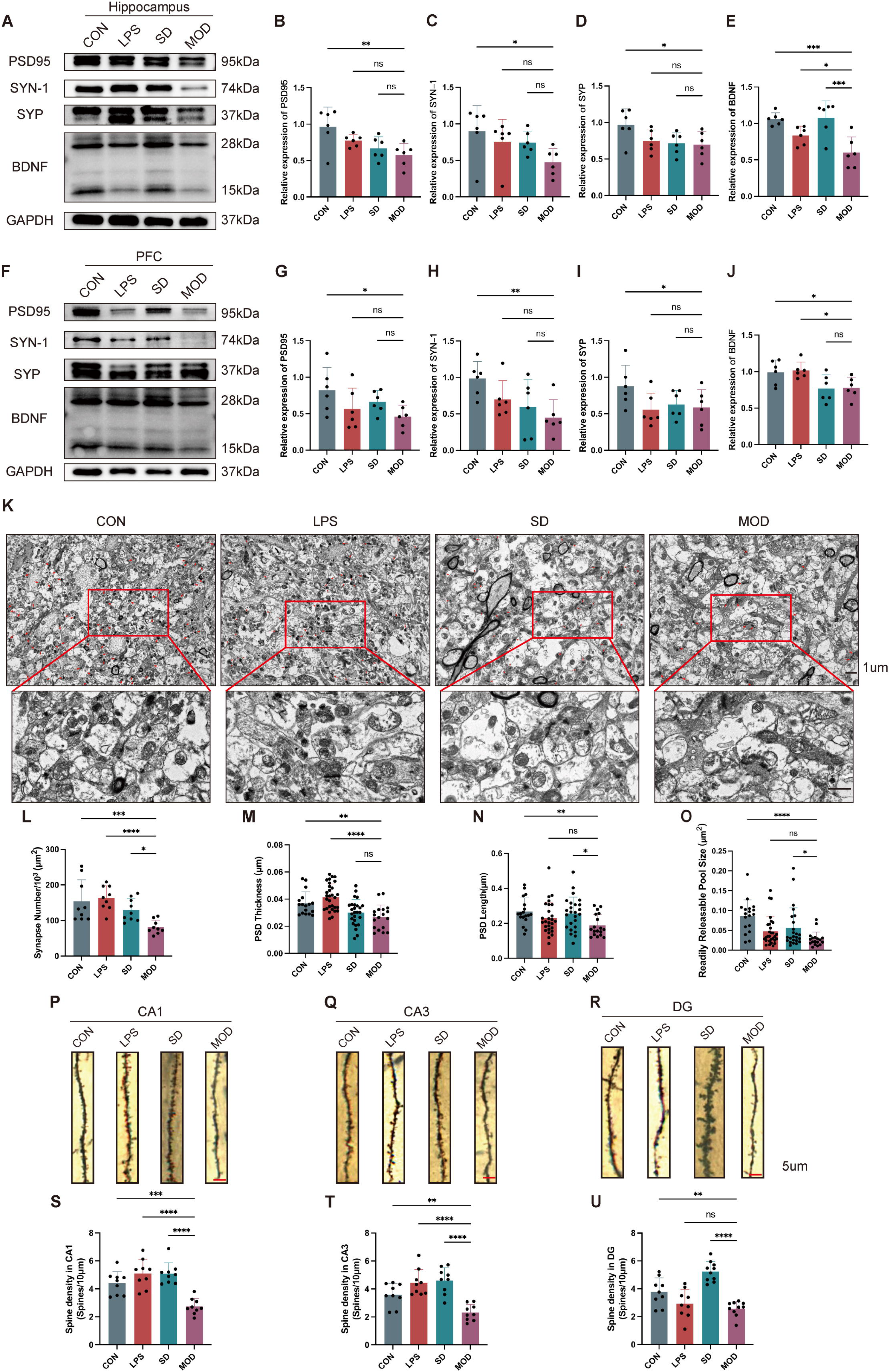
SD plus LPS treatment induces synapse dysfunction in both the hippocampus and PFC. Western blotting analysis showing reductions in PSD95, SYN-1, SYP, and BDNF in MOD hippocampus (A) and PFC (F). Quantification of the PSD95/GAPDH, SYN-1/GAPDH, SYP/GAPDH, and BDNF/GAPDH ratios (n = 6) in hippocampus (B-E) and PFC (G-J) tissue. Representative TEM images of synapses of hippocampal neurons in the indicated groups. Scale bars, 1 μm (black) (K). Quantitation of synapse density (n = 9) (L). PSD thickness (M), PSD length (N), and RRP size (O) in neurons were evaluated in the indicated groups (CON, n = 17; LPS, n = 30; SD, n = 26; MOD, n = 18). Representative Golgi-Cox staining of dendritic spines in CA1 (P), CA3 (Q), and DG (R) in the indicated groups. Scale bars, 5 μm (red). Quantitation of dendritic spine density in different hippocampal regions as mentioned above (n = 9) (S-U). Data are expressed as mean ± SD. \**p* < 0.05, \*\**p* < 0.01, \*\*\**p* < 0.001, \*\*\*\**p* < 0.0001 by one-way ANOVA with Fisher’s LSD multiple-comparisons test.

In the fluoxetine rescue experiment, as expected, the fluoxetine group exhibited increased protein expression of PSD95, SYN, SYP, and BDNF in the hippocampus (*p* < 0.05). Furthermore, reversible alterations in synaptic ultra-structures and dendritic spine morphology were observed within the fluoxetine group. However, no similar changes were observed in the PFC (**Supplemental Fig. 6**).

### Validation of depression-related DEGs based on TWAS

A central goal of this study was to validate DEGs identified in the two-hit model (model group) using data derived from human genetics studies of brain tissues. Thirty-three genes overlapped by intersecting DEGs with genes associated with depression in TWAS (Benjamini-Hochberg correction *p* < 0.05), with 29 found in the hippocampus and four in the PFC (**Supplemental Table 3**). By comparing Z-scores from TWAS with identified DEGs in transcriptomes analyses, we observed consistent directional changes in expression for 16 genes (**Fig. 6**). Interestingly, the significant TWAS gene set contained genes that regulate inflammatory and immune responses, exemplified by *TLR9* and *C4A*, and those implicated in the modulation of neurotransmitter release, like *CTTNBP2*. These findings are consistent with our experimental observations, reinforce the two-hit model, and suggest a fundamental connection between insomnia, inflammation, and depression.

**Fig. 6.**
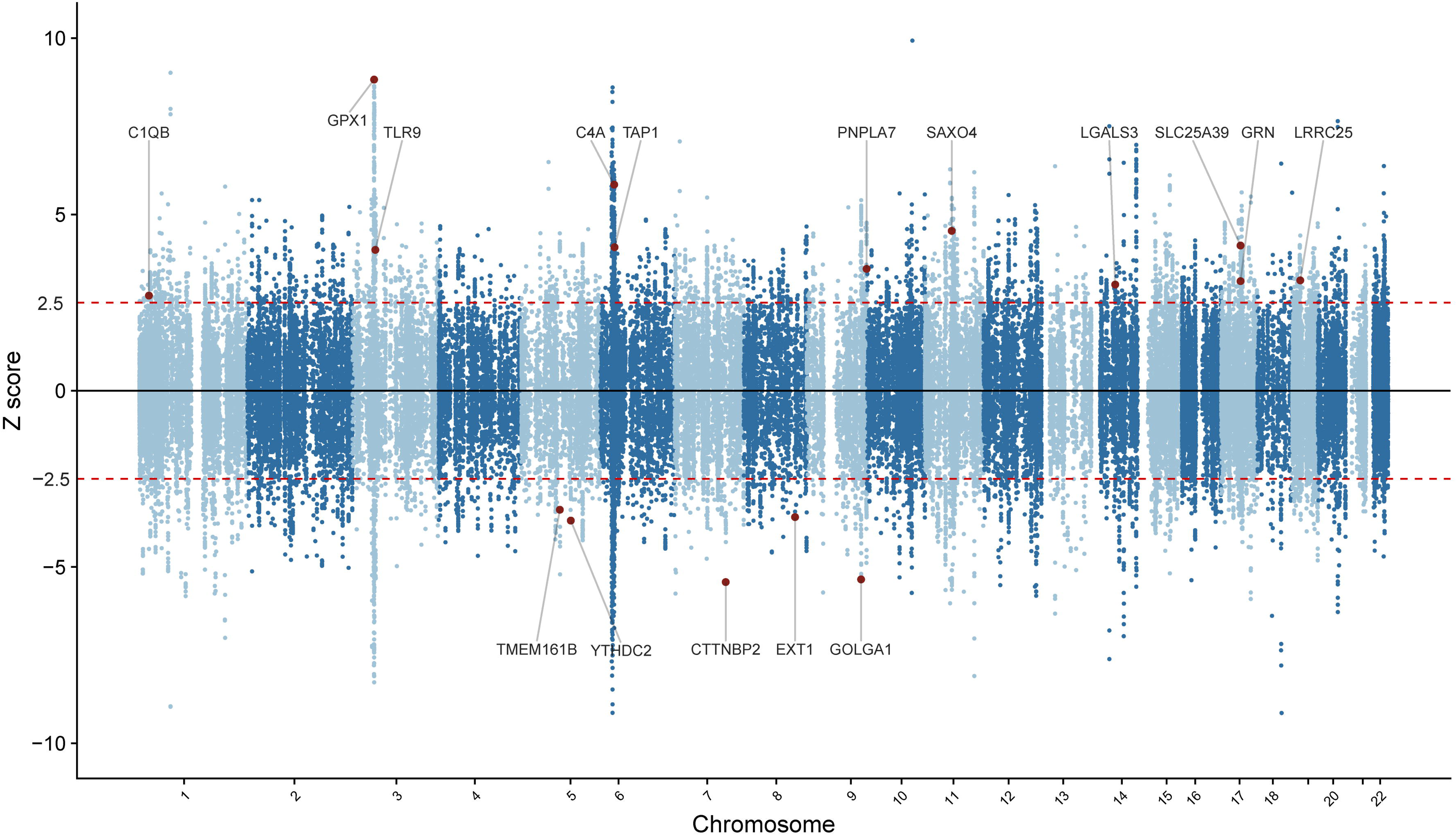
Manhattan-style Z-score plot illustrating validated gene associations with depression in the TWAS analysis. TWAS results confirm the differential gene expression model. Sixteen validated genes from TWAS results across 13 brain tissues based on depression GWAS are represented as red dots. The red dashed lines indicate the TWAS significance threshold following Benjamini and Hochberg correction (*FDR.p* < 0.05). Genes positioned in the upper portion of the plot indicate that increased expression correlates with increased depression risk, while genes in the lower portion demonstrate an inverse association, consistent with the direction of the differential gene expression model.

## DISCUSSION

In this study, based on the two-hit hypothesis [16], we devised a novel rodent model of depressive-like behaviors that used chronic SD to simulate insomnia and LPS treatment to simulate inflammation. The model was validated from the etiological, phenomenological, and therapeutic perspectives. Additionally, significant neuroinflammation and impaired synaptic plasticity were observed in the model group, which are consistent with observations in the clinical trials. Furthermore, validation of a subset of the DEGs identified in model rats through human TWAS analysis supports the two-hit model of depression development in humans.

A key criterion of efficacy of depression models is the assessment of depressive-like behaviors. We employed widely used depressive-like behavior tests to assess the models, with the results revealing that the model group exhibited typical depressive-like behaviors, consistent with findings from previous of depression models in rodents, such as unpredictable chronic mild stress, repeated restraint stress, and chronic social defeat stress [24–26].

Of note, after randomly assigning rats to different groups at baseline, we did not exclude any rats. Combined with the results of behavior tests, we believe that an advantage of this model is its high success rate and reliability. Moreover, fluoxetine rescued depressive-like behaviors, validating the model’s efficiency from a therapeutic perspective. Thus, this novel rodent model fulfilled the three criteria for a depressive-like rodent model: (a) face validity (symptomatic homology), (b) etiological validity (similar causative factors), and (c) pharmacological validity (reversal of depressive symptoms by antidepressants) [27, 28].

A useful animal model of depression should not only demonstrate depressive-like behaviors but also recapitulate some main pathophysiological mechanisms responsible for depression. We therefore tested whether the model demonstrated some biological alterations seen in patients with depression. To obtain a preliminary picture of the model’s impact on the brain, we selected two brain regions widely studied in depression research, the hippocampus and the PFC, and conducted transcriptomic analysis.

Our results revealed significant enrichment of DEGs in signaling pathways such as “inflammatory and immune response”, “postsynaptic density”, “dendritic spine”, “regulation of synaptic plasticity”, and “postsynapse organization” in the model group (**Fig. 2**). These pathways are crucial in neurotransmission and neurodevelopment and correspond to impairments found in human studies [29, 30]. To ensure the robustness of our transcriptomics findings, we further validated neuroinflammation and impaired synaptic plasticity at the cellular and protein levels.

Neuroinflammation is one of the most reproduced phenomena in patients with depression. A meta-analysis summarizing central inflammatory markers associated with patients with depression suggested significantly elevated levels of proinflammatory cytokines in the cerebrospinal fluid [31]. Our two previous clinical trials of blood astrocyte-derived extracellular vesicles also provided direct *in vivo* evidence of neuroinflammation in patients with depression [32, 33]. Western blotting validation also revealed upregulation of neuroinflammatory proteins, while immunofluorescence indicated activation of microglia and astrocytes, suggesting signs of neuroinflammation (**Fig. 4**). Similar results have also been observed in humans [34–36]. Based on the transcriptomic data and comprehensive validations, we believe that this model effectively simulates neuroinflammation-related mechanisms of depression.

Our results also illustrate the capacity of the two-hit model to replicate alterations in synaptic plasticity, a frequently noted and highly replicable phenomenon in patients with depression [37]. Transcriptomic analysis revealed impaired synaptic plasticity in both the hippocampus and the PFC. This finding was further corroborated by western blotting, which demonstrated a decrease in the expression of synaptic marker proteins. Additionally, TEM and Golgi-Cox staining revealed a reduction in the number of synapses and dendritic spines, respectively. Alterations in the density, number, and morphology of synapses and dendritic spines have been observed in both depressive-like animal models and patients with depression [38]. Imaging and postmortem studies have also suggested that synapse number and function are decreased in patients with major depressive disorder [39]. Additionally, the number and efficiency of synapses are also influenced by a variety of factors related to depression and other mood disorders, such as neurotrophic factors and inflammatory cytokines [40, 41]. In summary, both clinical and basic research highlight a pivotal role for synaptic plasticity in the neurobiology of depression [37, 42, 43], further supported by our findings within the two-hit model framework.

The pattern observed in the PFC was similar to that observed in the hippocampus, albeit with subtle differences. Compared with controls, both the LPS and SD groups showed a reduction in the expression of synaptic marker proteins within the PFC. This phenomenon is thought to arise from a dominant cortical involvement induced by LPS treatment and SD, with the hippocampus affected to a lesser extent [44, 45].

Although rats in the LPS group also displayed depressive-like behaviors, consistent with previous reports [46, 47], we chose SD plus LPS as the model treatment. This was for two reasons. First, based on clinical evidence, we used SD plus LPS to simulate the scenario seen in humans [15, 48]. Second, in our study, the impact of SD plus LPS on the brain seemed different to the synapse structure observed with SD or LPS alone. Compared with controls, no significant changes were observed in the number of synapses, PSD thickness, or dendritic spine density in the hippocampus of the LPS group.

Interestingly, according to TEM and Golgi-Cox staining, there was a significant increase in the density of hippocampal dendritic spines in the SD group compared with control and model animals (**Fig. 5**). Previous human and animal studies have demonstrated that wakefulness increases synaptic strength in the cerebral cortex, whereas sleep can reduce cortical synapses to restore synaptic balance. Many dendritic branches show a net reduction in spines after sleep, with the remaining synapses generally being smaller [49, 50]. This phenomenon is known as the synaptic homeostasis hypothesis [51, 52], and it effectively explains the changes in synaptic function and structure observed in the SD group. Another interesting hint is that the combination of SD and inflammation, the “two hits”, can lead to structural and functional impairments in synapses in the hippocampus, consistent with clinical findings [51, 53, 54]. This reduction may be related to sleep pressure [55] and inflammation. First, chronic SD results in a net increase in synaptic strength to saturation, with a subsequent sudden increase in systemic inflammation exerting a dual impact of extensive synaptic transmission disruption and synaptic damage, ultimately leading to decreased synaptic plasticity [51, 56]. This interesting possibility now requires further exploration.

Another key criterion for a valid depression model is that model animals can be rescued by classical antidepressants. In our study, model rats demonstrated a positive response to fluoxetine treatment. More importantly, following treatment, there were improvements in both behavioral outcomes and neuroinflammation and synaptic plasticity. Fluoxetine significantly downregulated neuroinflammatory marker proteins, while synapse-related proteins were upregulated. Immunofluorescence analysis revealed that fluoxetine rescued alterations in GFAP^+^ and IBA-1^+^ cells and reversed changes in synaptic microstructure and dendritic spine morphology. A meta-analysis suggested that fluoxetine modulates the recovery of neurotransmission and neuroinflammatory pathways in human depression [57]. Furthermore, Alboni et al. reported that fluoxetine may also enhance neuroplasticity in depression [58]. These findings are consistent with our observations in fluoxetine rescue experiments. However, the results of the rescue experiment indicated that fluoxetine treatment did not ameliorate synaptic dysfunction in the PFC of the MOD rats. It was likely due to the more severe synaptic dysfunction inflicted on the PFC by the combined LPS and SD treatment compared to the hippocampus, which is mentioned above. A three-week fluoxetine treatment may not be sufficient to alleviate synaptic dysfunction in the PFC, suggesting that a longer treatment duration may be required. This necessitates further exploration in future studies.

If our model is an effective model of human disease, we would expect it to simulate transcriptional level changes seen in the brains of patients with depression. We therefore conducted TWAS analyses of profiles obtained from human brain tissues corroborating the two-hit model of depression. Of 33 validated genes, 16 exhibited consistent directional changes in expression, with five genes having negative Z-scores and 11 positive Z-scores in the TWAS analysis. Subsequent investigation of these depression risk genes revealed that five (*TLR9*, *C4A*, *LGALS3*, *C1QB*, and *GRN*) are implicated in the immune and inflammatory responses, while three (*GRN*, *C1QB*, and *CTTNBP2*) are involved in synaptic pathways.

There is existing evidence to support the involvement of *TLR9* and *C4A* in the pathogenesis of depression. *TLR9*, a member of the toll-like receptor (TLR) family, activates proinflammatory cytokine production. Pre-clinical and clinical studies have demonstrated that TLR expression and TLR signaling regulators are associated with depression [59]. Additionally, TLR also plays a role in innate immunity as measured in the postmortem brains of patients with depression and suicide [60]. Dysregulation of *C4A* expression or activity has been implicated in the perpetuation of neuroinflammation, with genetic studies associating *C4A* polymorphisms with an increased susceptibility to depression [61, 62]. Future studies could explore their potential as diagnostic biomarkers and therapeutic targets in depression. *CTTNBP2* plays a crucial role in regulating dendritic spine formation, modulating synaptic signaling through its effects on the protein phosphatase 2A (PP2A) complex, potentially affecting the specificity and efficiency of synaptic transmission [63]. It also regulates AMPA receptor activity by influencing the trafficking machinery, thereby impacting synaptic strength and plasticity [64]. The validation of these genes in human genetics studies further supports the two-hit model and reinforces the fundamental role of neuroinflammation and synaptic dysfunction in depression.

This study has several limitations. First, depression is a complex disease with potentially multiple etiologies, and this model may not fully simulate all of these etiologies. This model might need to be integrated with other existing models to provide further insights into depression. Second, in our analysis, we selected two representative brain regions for analysis, the hippocampus and PFC, but depression may affect other brain areas. Future studies could apply advanced techniques such as single-cell sequencing, spatial transcriptomics, and neural circuit techniques across the brain to further explore the model. Third, the transcriptomic analysis yielded other potentially important pathways, such as mitochondrial respiratory chain and learning and memory, but owing to space constraints, we focused on validating neuroinflammation and synaptic plasticity as they are among the most reproducible phenomena in clinical depression. However, this does not diminish the importance of other findings in the model group, which require further exploration. Lastly, the TWAS results need further validation, as TWAS power can be influenced by the quality of gene expression (sample sizes, coverage of eQTLs in the test dataset, *etc.*)

In conclusion, here we introduce a novel two-hit model of depression induced by SD and LPS that effectively replicates depressive-like behaviors in rodents. This novel rat model could be a valuable research tool for further investigation of the roles of insomnia and inflammation in the pathophysiology of depression.

### Data availability

All data generated or analyzed during this study are either included in this published article or are available from the corresponding author upon reasonable request.

## Supporting information

sTable 1

sTable 2

sTable 3

## Acknowledgements

The authors are very grateful for the data support provided by the GTEx database and iPSYCH website. We gratefully acknowledge the contributions of Dr. Wei Wang, and Dr. Simeng Ma from Renmin Hospital of Wuhan University, as well as Dr. Zhiyu Luo from Wuhan First Hospital, to our work.

## Author contributions

**Junhua Mei**: Writing – original draft, Project administration, Funding acquisition, Methodology. **Xinhua Song**: Methodology, Validation, Construction of the two-hit rodent model, Basic Experiment, Formal analysis, Writing – original draft, Visualization. **Ying Wang**: Methodology, Formal analysis, Writing – original draft, Visualization. **Honggang Lyu**: Methodology, Formal analysis, Writing – original draft, Visualization. **Guang Wang**, **Chao Chen**, and **Honghan Zhang**: Validation, Construction of the two-hit model. **Chao Wang**: Methodology, Writing – review and editing. **Xin-hui Xie**: Conceptualization, Methodology, Formal analysis, Writing – review and editing, Supervision. **Guohua Chen** and **Zhongchun Liu**: Funding acquisition, Supervision.

