## Supplementary file 1 for "A novel two-hit insomnia and inflammation rodent model of depressive-like behaviors"

**Behavioral assessment**

**Sucrose preference test**

The SPT is a reward-based assessment used to measure anhedonia. In this test, rats were provided with two pre-weighed bottles: one containing a 2.5% sucrose water solution and the other filled with plain water. The rats were allowed to choose between these two options for 12 hours. The total volume of pure water and sucrose water consumed was calculated separately.

**Forced Swimming Test**

The FST is designed to simulate the behavioral response of rats when they are in a stressed state. The forced swimming bucket is thoroughly cleaned to remove any animal odors. In a cylindrical container with a total volume of 2500 mL, a height of 20 cm, and a diameter of 14 cm, the container is filled with 10 cm of water at a temperature of 25°C. The rats are placed in the cylinder, and the time spent in forced swimming is recorded for 6 minutes.

**Western blotting**

Total protein was extracted from rats' HIP and PFC using a lysis buffer (KeyGEN, Wuhan, China). After quantification with the BCA kit (KeyGEN, Nanjing, China), we loaded the protein samples on sodium dodecyl sulfate-polyacrylamide gel and transferred them to polyvinylidene difluoride (PVDF) membranes. The primary antibodies and glyceraldehyde-3-phosphate dehydrogenase (GAPDH) were applied overnight at 4℃ after incubation with 5% skim milk for one hour. Then, the secondary antibody (KeyGEN, Nanjing, China) was coupled with horseradish peroxidase to coat the membrane for one hour at room temperature. Finally, we visualized the proteins with a chemiluminescence kit (Millipore, Billerica, MA, USA)

**Immunofluorescence (IF)**

Brain tissues were fixed, embedded, and sectioned into 4 μm thick slices. After dewaxing and rehydration, antigen retrieval was performed. The sections were then blocked with 10% BSA and incubated overnight with primary antibodies. Following a 1-hour incubation with secondary antibodies at room temperature, the sections were stained with DAPI for 5 minutes. Fluorescence images were captured using the microscope (Olympus BX51).

**Transmission electron microscopy (TEM)**

Brain tissues were carefully dissected into 1 mm³ segments and then fixed in 2.5% glutaraldehyde for 4 hours, followed by 1% osmium acid for 2 hours. After ethanol dehydration, these segments were immersed in a semi-epoxy-propane mixture overnight, embedded in resin, and cut into 70 nm slices. The ultrathin slices were stained on copper grids with 4% uranyl acetate and 0.5% lead citrate. The morphology of hippocampal synapses was observed by TEM (Hitachi, HT7700).

**Golgi-Cox staining**

Hippocampal tissue blocks were fully immersed in Golgi dye solution (Servicebio, G1069) for 14 days, with the staining solution refreshed every 3 days. Following immersion in 80% glacial acetic acid, the blocks were dehydrated in 30% sucrose and then sectioned into 100 μm slices. After treatment with ammonia and acid-hardening fixing solution, the slices were sealed using glycerin gelatin.

**Supplemental Fig. 1**. Separate plots illustrating the distribution of PCA-based samples, organized by gene expression in hippocampus (A) and PFC (B). The Venn diagram shows the differential expressed genes among the MOD, LPS, and SD groups in hippocampus (C) and PFC (D).

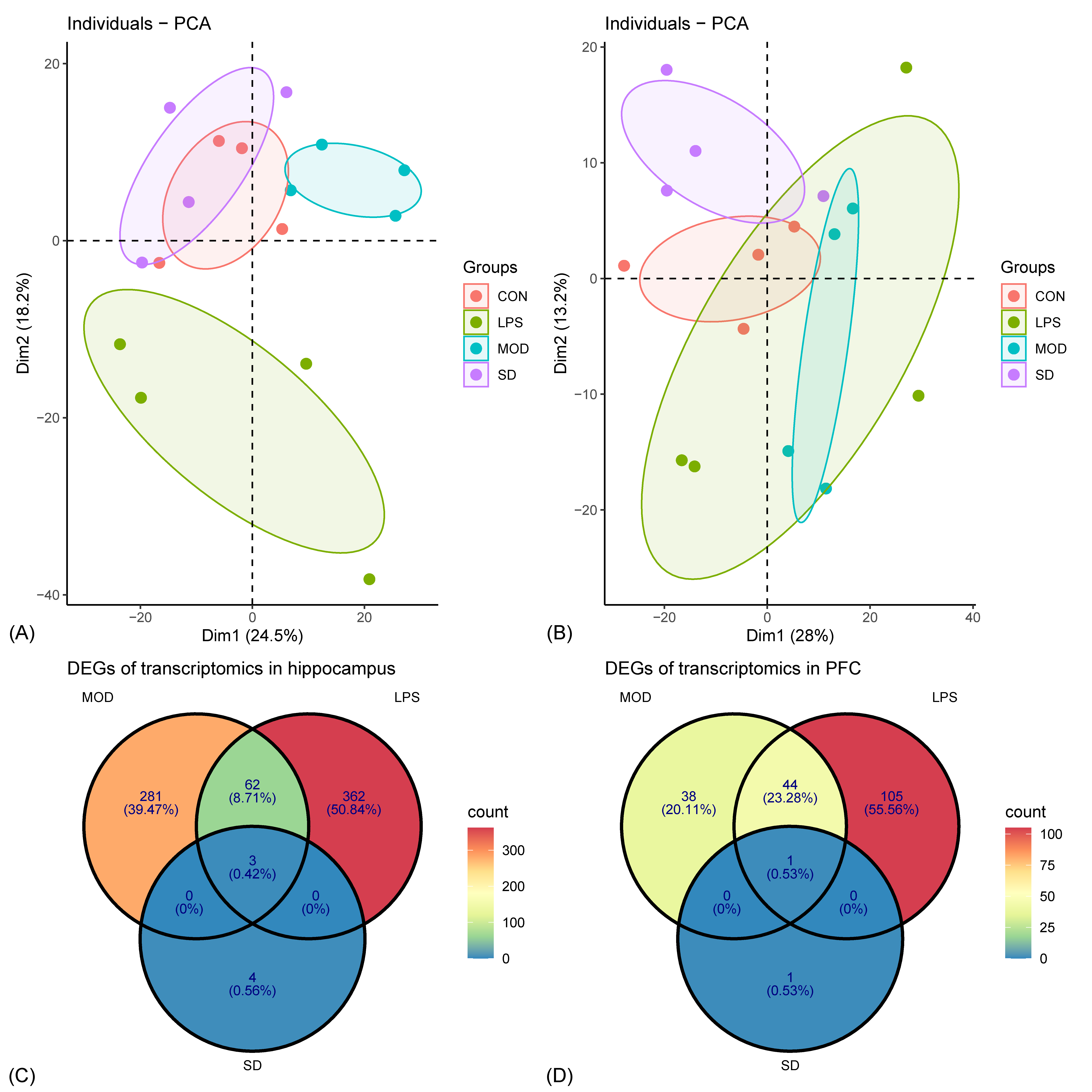

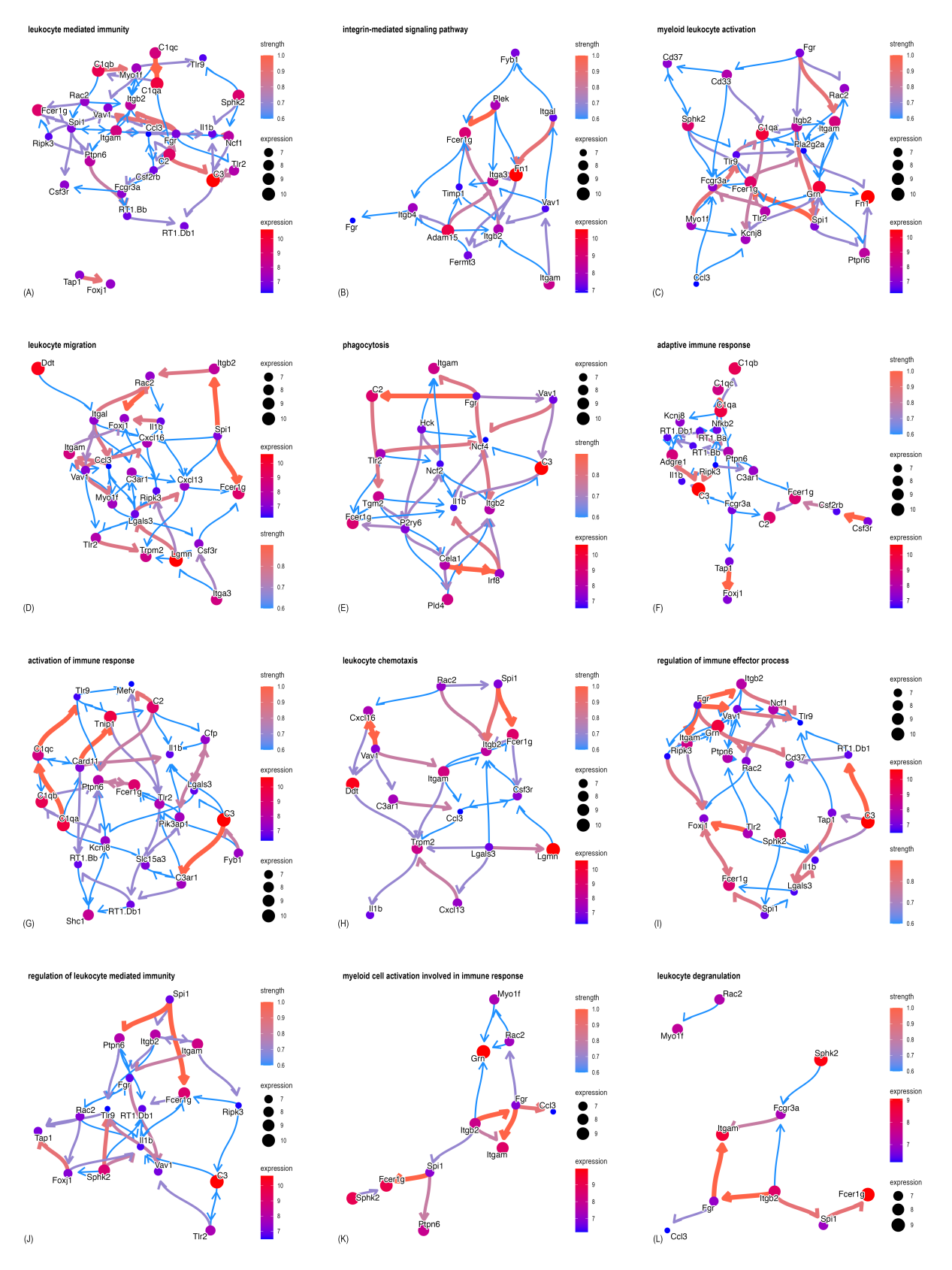
**Supplemental Fig. 2**. Perform GO enrichment analysis on the upregulated DEGs in the hippocampal MOD group, showing pathways related to immune-inflammatory response.

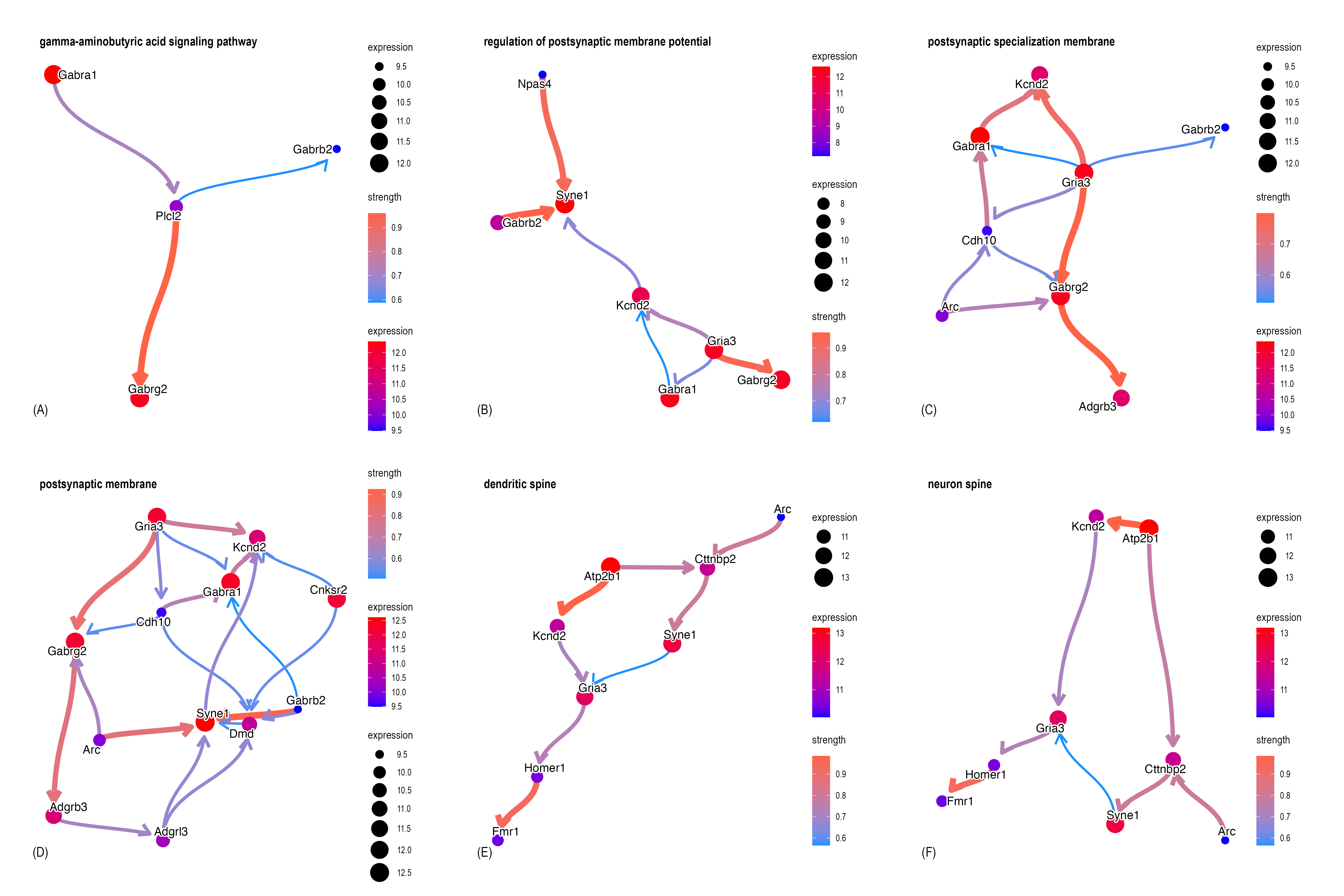
**Supplemental Fig. 3**. Enrichment analysis on the downregulated DEGs in the hippocampal MOD group, showing pathways related to synapses.

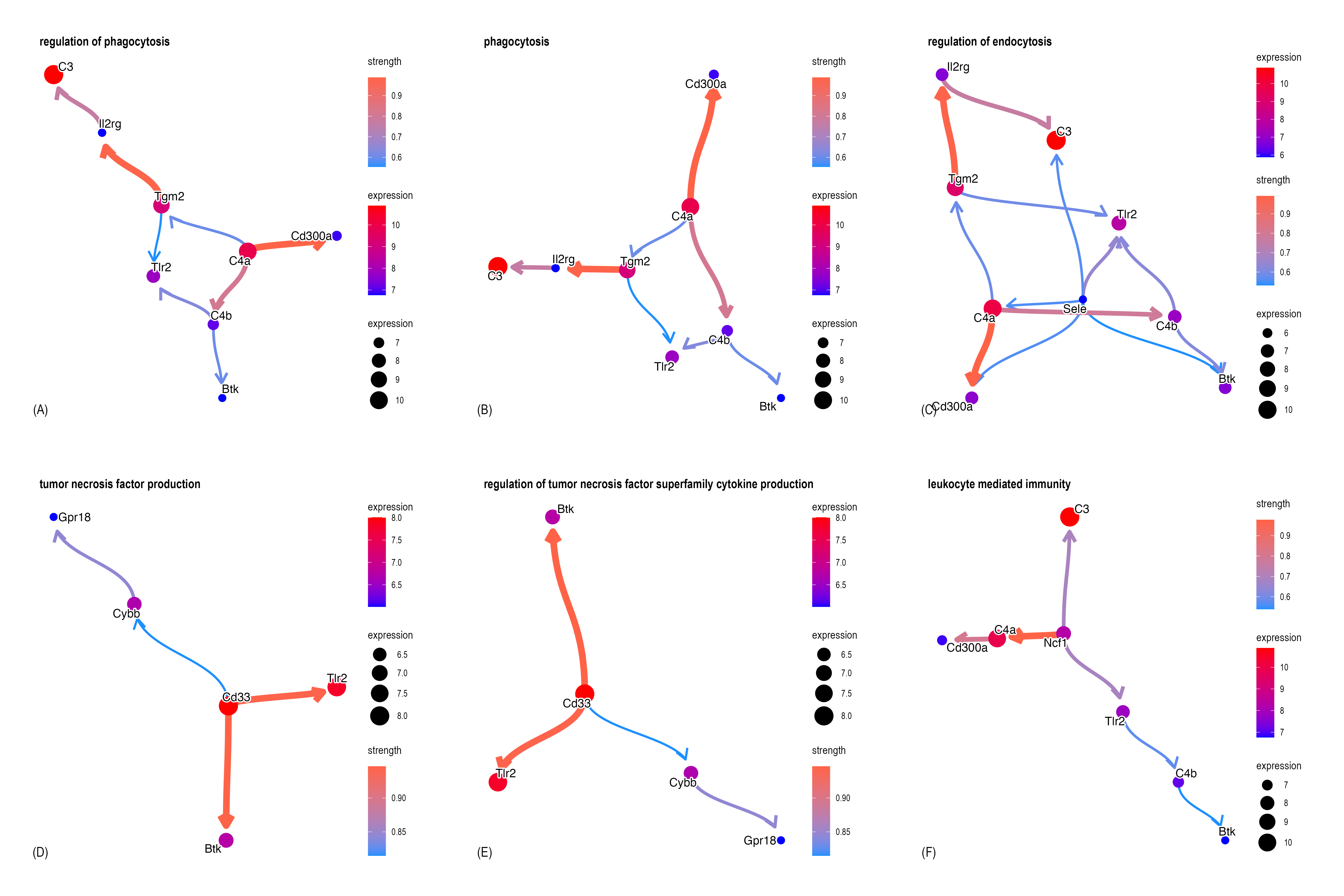
**Supplemental Fig. 4**. Enrichment analysis on the upregulated DEGs in the PFC MOD group, showing pathways related to immune-inflammatory response.

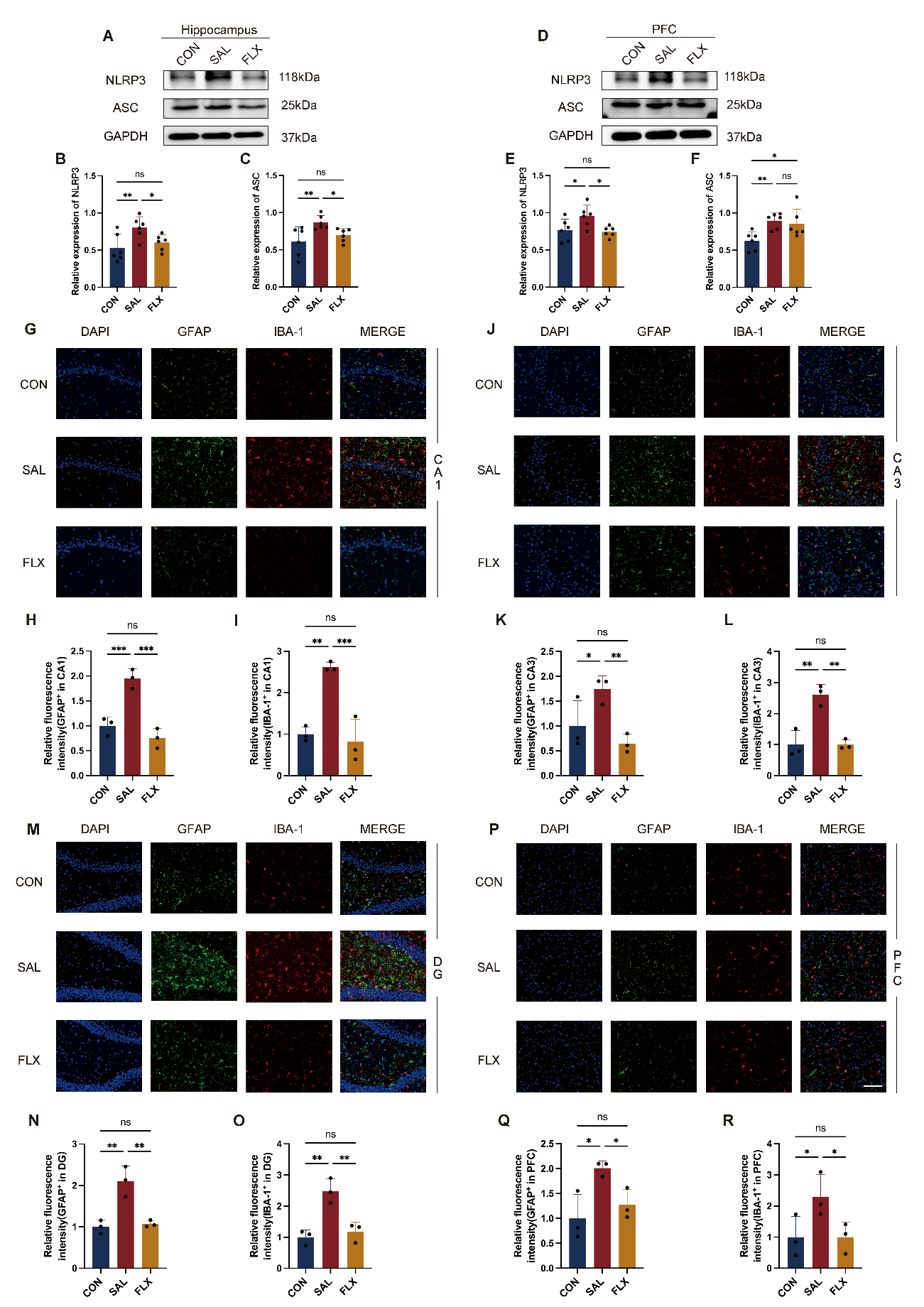
**Supplemental Fig. 5**. Fluxoxetine treatment could rescue neuroinflammation in both hippocampus and PFC which induced by sleep deprivation and LPS. Western blotting analysis showing the decrease of NLRP3, ASC in FLX hippocampus (A) and PFC (D). Quantification of the NLRP3/GAPDH, ASC/GAPDH ratio (n = 6) in hippocampus tissue (B, C) and PFC tissue (E, F). Representative immunofluorescence images of GFAP^+^ and IBA-1^+^ cells in CA1 region (G), CA3 region (J), DG region (M) and PFC region (P) in the indicated groups. Scale bars, 100 μm (white). Quantitation of the GFAP^+^ cells relative fluorescence intensity in the indicated groups (H, K, N, Q). Quantitation of the IBA-1+ cells relative fluorescence intensity in the indicated groups (I, L, O, R). Data were expressed as mean ± SD. **p* < 0.05, ***p* < 0.01, ****p* < 0.001, *****p* < 0.0001, by one-way ANOVA with Fisher's LSD muliple-comparisons test.

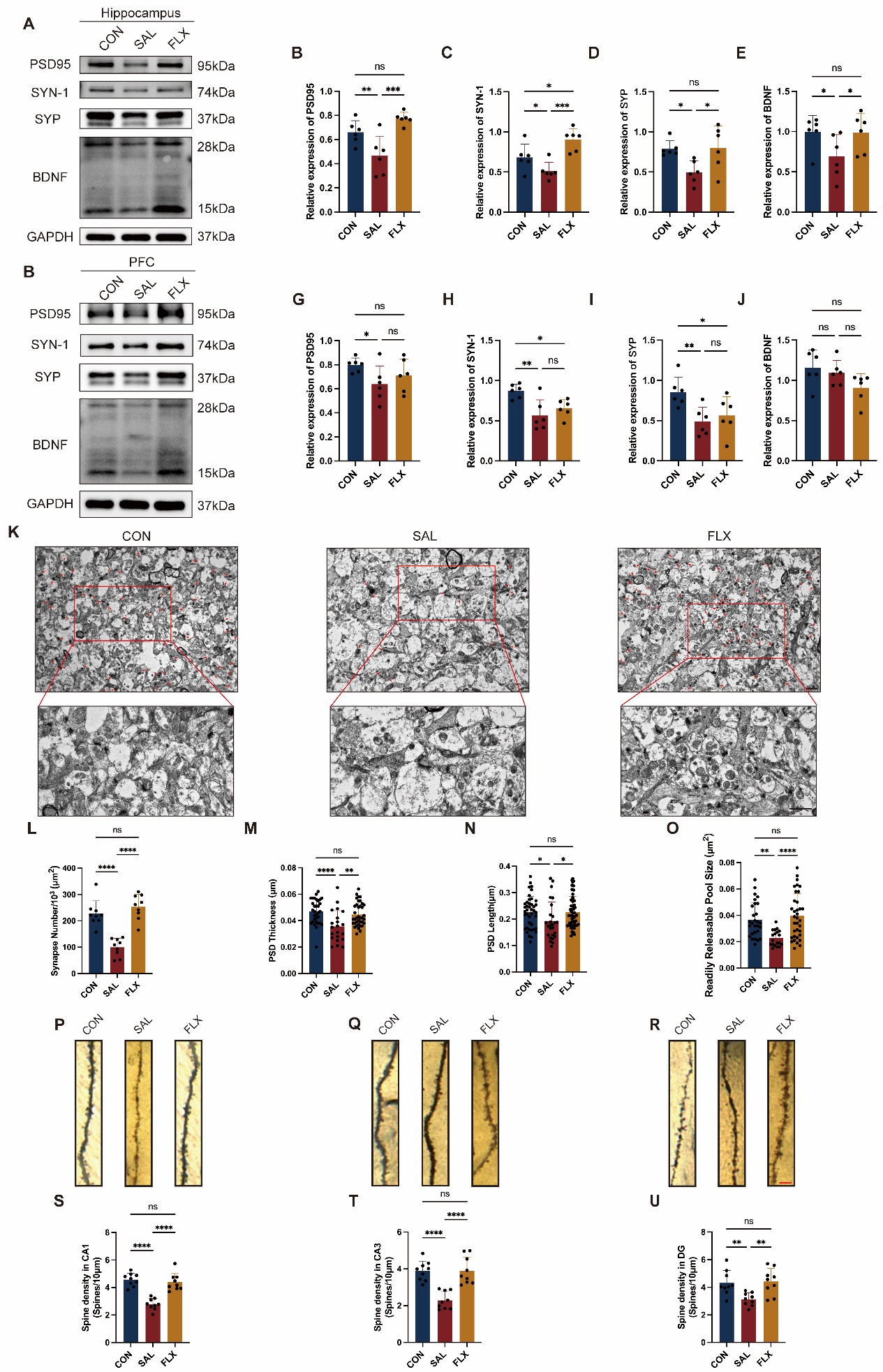

**Supplemental Fig. 6**. Fluxoxetine treatment could alleviate synapse dysfunction in hippocampus instead of PFC. Western blotting analysis showing the increase of PSD95, SYN-1, SYP and BDNF in FLX hippocampus (A) and PFC (F). Quantification of the PSD95/GAPDH, SYN-1/GAPDH, SYP/GAPDH and BDNF/GAPDH ratio (n = 6) in hippocampus tissue (B-E) and PFC tissue (G-J). Representative TEM images of synapses of hippocampus neurons in the indicated groups. Scale bars, 1 μm (black) (K). Quantitation of the synapse density (n = 9) (L). PSD thickness (M), PSD length (N) and RRP size (O) in the neurons were evaluated in the indicated groups. Representative Golgi-Cox staining images of the dendritic spine in the CA1 region (P), CA3 region (Q) and DG region (R) in the indicated groups. Scale bars, 5 μm (red). Quantitation of the dendritic spine density in different hippocampal regions as mentioned above (n = 9) (S-U). Data were expressed as mean ± SD. **p* < 0.05, ***p* < 0.01, ****p* < 0.001, *****p* < 0.0001, by one-way ANOVA with Fisher's LSD multiple-comparisons test.
